# Interactive web application for plotting personalized prognosis prediction curves in allogeneic hematopoietic cell transplantation using machine learning

**DOI:** 10.1101/2019.12.14.19014654

**Authors:** Hiroshi Okamura, Mika Nakamae, Shiro Koh, Satoru Nanno, Yasuhiro Nakashima, Hideo Koh, Takahiko Nakane, Asao Hirose, Masayuki Hino, Hirohisa Nakamae

**Author notes:** **Contact information for correspondence:** Hiroshi Okamura, M.D., Ph.D. Hematology, Graduate School of Medicine, Osaka City University, 1-4-3 Asahi-machi, Abeno-ku, Osaka 545-8585, Japan, **E-mail:**. **AUTHORSHIP:** H.O. participated in the conception and design of the study, analysis of data, and interpretation of data and drafted the paper. M.N. participated in the construction of database and interpretation of data and revised the paper. A.H. participated in the construction of database and revised the paper. S.K., S.N., Y.N, H.K, T.N, M.H., and H.N. participated in the acquisition of data and interpretation of data and revised the paper. All authors critically reviewed the paper.

## Abstract

**Background:** Allogeneic hematopoietic cell transplantation (allo-HCT) is a curative treatment option for malignant hematological disorders. Transplant clinicians estimate patient-specific prognosis empirically in clinical practice based on previous studies on similar patients. However, this approach does not provide objective data. The present study primarily aimed to develop a tool capable of providing accurate personalized prognosis prediction after allo-HCT in an objective manner.

**Methods:** We developed an interactive web application tool with a graphical user interface capable of plotting the personalized survival and cumulative incidence prediction curves after allo-HCT adjusted by eight patient-specific factors, which are known as prognostic predictors, and assessed their predictive performances. A random survival forest model using the data of patients who underwent allo-HCT at our institution was applied to develop this application.

**Results:** We succeeded in showing the personalized prognosis prediction curves of 1-year overall survival (OS), progression-free survival (PFS), relapse/progression, and non-relapse mortality (NRM) interactively using our web application (https://predicted-os-after-transplantation.shinyapps.io/RSF_model/). To assess its predictive performance, the entire cohort (363 cases) was split into a training cohort (70%) to develop the predictive model and test cohort (30%) to confirm its performance time-sequentially. The areas under the receiver-operating characteristic curves for 1-year OS, PFS, relapse/progression, and NRM in test cohort were 0.70, 0.72, 0.73, and 0.77, respectively.

**Conclusions:** The new web application could allow transplant clinicians to inform a new allo-HCT candidate of the objective personalized prognosis prediction and facilitate decision-making.

## INTRODUCTION

Allogeneic hematopoietic cell transplantation (allo-HCT) is an advanced therapeutic intervention required for high-risk malignant hematological disorders.^1^ Various complex and intertwined pre-transplant factors affect patient prognosis after allo-HCT.^2–5^ Transplant clinicians need to estimate individual prognosis after allo-HCT in clinical practice by combining not only certain indices that can stratify the prognosis after allo-HCT, such as refined Disease Risk Index (DRI-R), EBMT risk score, and Hematopoietic Cell Transplantation Comorbidity Index (HCT-CI), but also individual factors (e.g., age, performance status [PS], disease status, or the number of transplantations) and transplant procedures (e.g., donor source or conditioning regimen).^6–8^ However, the predictive capacities of these prognostic indices remain suboptimal.^2^ Furthermore, prognosis prediction based on the empirical integration of multiple patient-specific factors by the clinician is not always objective due to the complexity of allo-HCT treatment.

A random survival forest (RSF) is an ensemble machine learning method for right-censored time-to-event data analysis. Recently, RSF has become increasingly popular for survival data analysis in various medical fields because of its high-precision predictions.^9–11^ It is possible to develop a prognosis prediction model integrating multiple factors to compute patient-specific prognosis prediction curves such as survival or cumulative incidence adjusted for individual factors of a new patient using RSF.^12^ Although the most popular model for survival analysis is the Cox proportional hazard (Cox PH) regression, RSF has some advantages over the Cox PH model. First, RSF can easily deal with non-linear effects, correlated parameters, and variable interaction. Second, while the Cox PH model may be limited due to the proportional hazards assumption, RSF is non-parametric and completely independent of model assumptions.^11,13^ Therefore, survival prediction analysis by RSF may provide novel insights into the allo-HCT field.

Although objective and accurate patient-specific prognosis prediction using a machine learning model can be quite informative for clinicians and patients in clinical decision-making, it has not been applied in a clinical setting. A user-friendly tool for constructing accurate personalized prognosis prediction curves after allo-HCT is lacking. Such a tool could allow transplant clinicians to inform patients of patient-specific prognosis prediction objectively derived from past patient data, and build an optimal personalized treatment strategy.

Here, we developed a web application tool with a graphical user interface (GUI) capable of plotting the personalized survival and cumulative incidence prediction curves after allo-HCT adjusted by eight patient-specific factors, which are known as prognostic predictors, using RSF and assessed their predictive performances.

## MATERIALS AND METHODS

### Patients and Definition

This study, comprising retrospective prognostic modeling, analyzed a group of consecutive patients who underwent allo-HCT at our institution between January 2008 and November 2017. All pre-transplant factors were recorded in the medical chart within 28 days prior to conditioning. The conditioning regimen containing either total body irradiation in fractionated doses greater than 8 Gy, an oral busulfan dose of 9 mg/kg or more, an intravenous busulfan dose of 7.2 mg/kg or more, or a melphalan dose of 140 mg/m^2^ or more was defined as myeloablative in line with a previous report.^14^ Human leukocyte antigen (HLA) compatibility was defined by DNA typing for HLA-A, HLA-B, HLA-C, and HLA-DR. This study was approved by the Osaka City University Hospital Certified Review Board. The context of this study was officially disclosed to the public through the posting of a notice at Osaka City University hospital and on the website of Hematology, Graduate School of Medicine, Osaka City University, according to the ethical guidelines for epidemiological research compiled by the Ministry of Education, Culture, Sports, Science and Technology, and the Ministry of Health, Labour and Welfare in Japan.

### Supportive Care

Prophylactic antibiotics, either levofloxacin or polymyxin B oral tablets, an antifungal agent, and acyclovir were routinely administered from the start of conditioning. Trimethoprim-sulfamethoxazole was also administered from the start of conditioning to 2 days prior to allo-HCT and after neutrophil engraftment to prevent *Pneumocystis jirovecii*-induced pneumonia. Granulocyte colony-stimulating factor treatment was generally initiated from the day following HCT to neutrophil engraftment. Ursodeoxycholic acid was administered from the start of conditioning to prevent sinusoidal obstruction syndrome.^15^

### Predictors and Outcomes

Eight pre-transplant factors known as prognostic predictors for allo-HCT, including recipients’ age, DRI-R, HCT-CI, PS, donor source (related bone marrow [rBM], related peripheral blood [rPB], unrelated bone marrow [uBM], cord blood [CB], or haploidentical peripheral blood [haplo-PB]), HLA compatibility, conditioning intensity (myeloablative conditioning [MAC] or reduced intensity conditioning [RIC]), and the number of allo-HCT were used as predictive variables.^2–5^

The predictive objectives were prediction of 1-year overall survival (OS), 1-year progression-free survival (PFS), and 1-year cumulative incidences of relapse/progression and non-relapse mortality (NRM). An OS event was defined as death from any cause. PFS was defined as survival without relapse/progression. NRM was defined as death without relapse/progression. We considered cases that never achieved complete remission after allo-HCT as relapse/progression on day 1 after allo-HCT. Relapse/progression and NRM were treated as competing events with one another. The probabilities of OS and PFS were calculated using the Kaplan–Meier method. Cumulative incidences of relapse/progression and NRM were calculated using Gray’s method.

### Statistical Analysis

The following is a summary of the analysis outline: (i) preprocessing data quality assurance, (ii) development of a web application tool, and (iii) assessment of the predictive performance for OS, PFS, relapse/progression, and NRM in accordance with the Type 2b (nonrandom split-sample development and validation) of prediction model studies covered by the Transparent Reporting of a prediction model for Individual Prognosis Or Diagnosis (TRIPOD) statement.^16^ We had no missing data. The number of trees to develop the RSF model was set as 500. A log-rank splitting rule was used to grow survival trees of a forest.

We used R package randomForestSRC for analysis of the RSF model, R package timeROC for calculating AUCs, and R package shiny/R for development of the web application.^17,18^ All analyses were performed using R version 3.5.1. We adhere to the TRIPOD statement.^16^

### Performance Assessment of Predictive Model

All predictive performances were calculated using the time-dependent area under the receiver-operating characteristic curve (AUC).^18^ First, the entire cohort was split into a training cohort (70%) and test cohort (30%) time-sequentially. Second, the optimal number of variables randomly selected as candidates for splitting a node, which is a hyperparameter in the RSF model, for each outcome was tuned using 5-fold cross-validation method in the training cohort.^19^ It was defined as the one with the highest AUC value. Third, we assessed the predictive performances of the RSF, Cox PH model, DRI-R, and HCT-CI for 1-year OS and PFS, and of RSF, DRI-R, and HCT-CI for 1-year relapse/progression and NRM in test cohort. We plotted time-sequential AUCs per month from 3 to 12 months after allo-HCT in the test cohort to assess the continuous predictive performances in each prognosis prediction curve. The schema of the assessment method for these predictive models is shown in Figure S1.

### Development of a Web Application Tool

We developed a web application tool capable of plotting the personalized prognosis prediction curves of 1-year overall survival (OS), 1-year progression-free survival (PFS), and 1-year cumulative incidences of relapse/progression and non-relapse mortality (NRM) adjusted by eight patient-specific factors of a new allo-HCT candidate, using RSF predictive model trained by the entire cohort.

## RESULTS

### Patients and Transplantations

Three hundred and eighty-four patients underwent allo-HCT. One patient who underwent transplant from unrelated peripheral blood stem cells was excluded due to insufficient sample for analysis. Since DRI-R cannot be scored when there is benign or rare disease, 21 patients were also excluded (aplastic anemia, 11; Idiopathic cytopenia of undetermined significance, 1; chronic active Epstein–Barr virus disease, 4; plasma cell leukemia, 1; chronic myelomonocytic leukemia, 2; myelodysplastic/myeloproliferative neoplasms, unclassifiable, 1). In total, 363 patients were analyzed in this study. The clinical characteristics and outcomes in the training cohort (254/363), test cohort (109/363), and entire cohort are listed in Table 1.

**Table 1.**
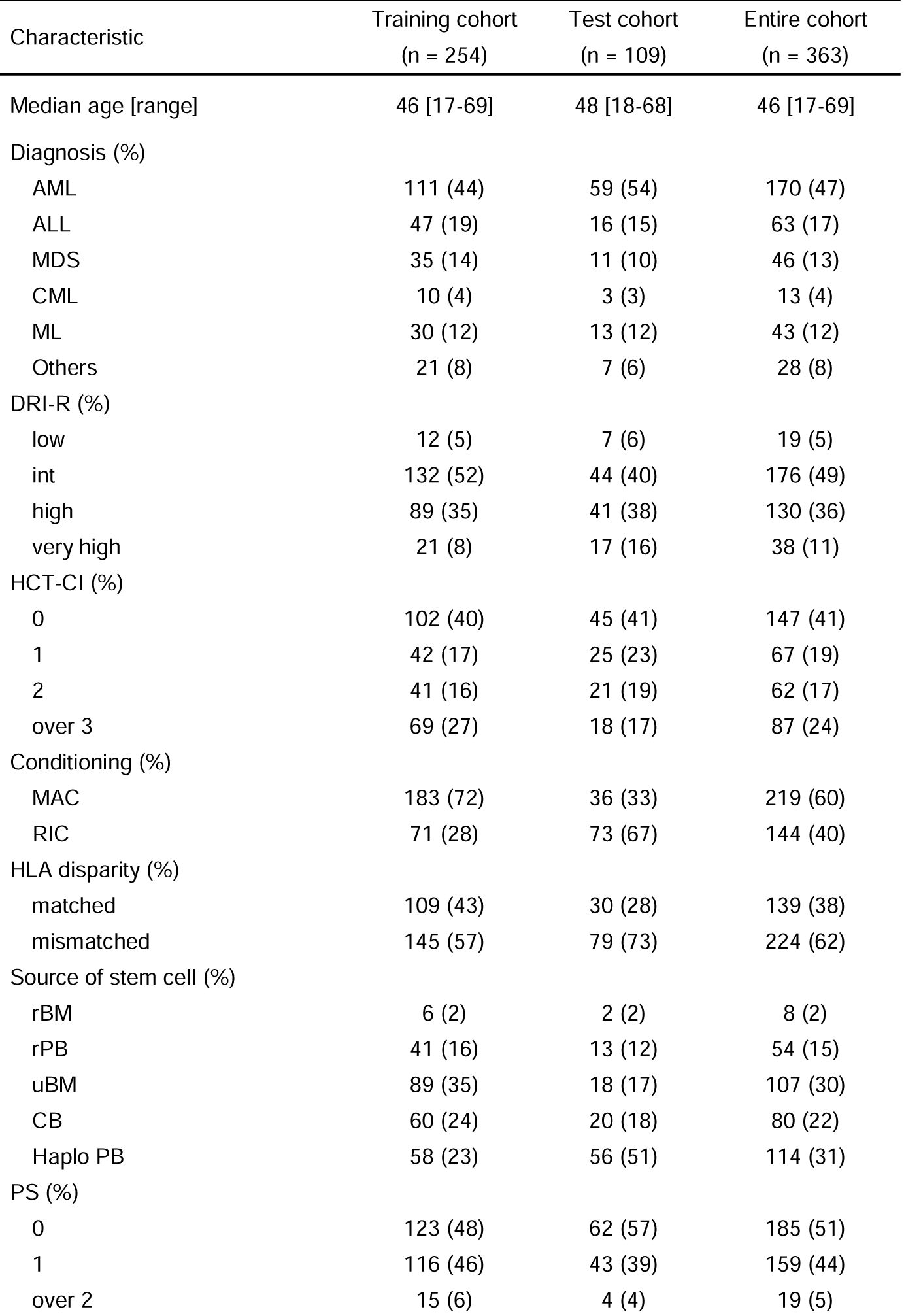
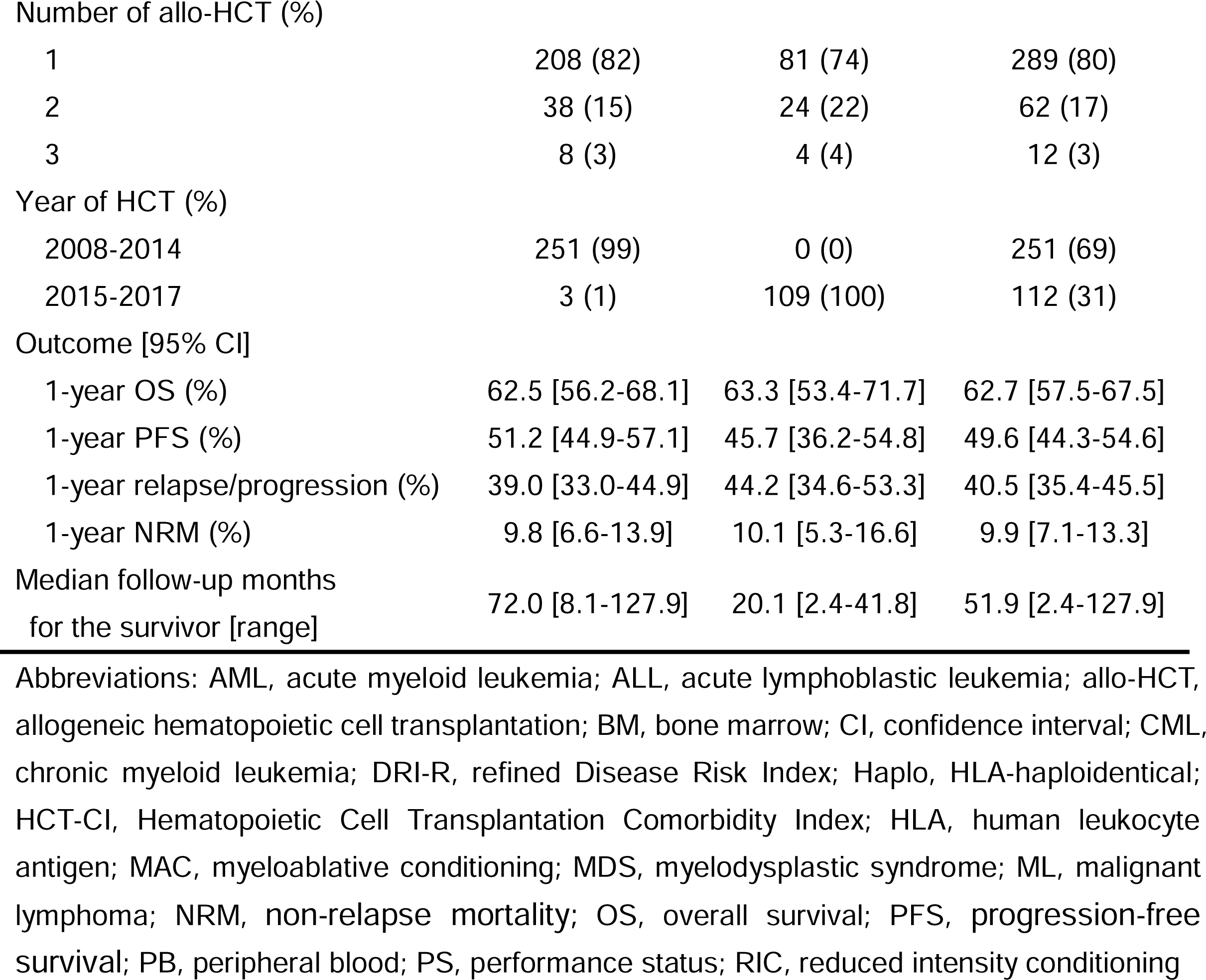
Patient Characteristics.

### Interactive Web Application for Plotting Personalized Prognosis Prediction Curves

The interactive web application that shows the personalized 1-year prognosis prediction curves after allo-HCT is shown in Figure 1. Users choose an outcome they intend to predict among OS, PFS, relapse/progression, or NRM in the select box at left-top (a). Next, after users set eight patient-specific factors in the left side-panel (b) and click the “prediction” button (c), the personalized 1-year prognosis prediction curve is displayed in the main panel (d). For example, Figure 1 shows the personalized 1-year OS prediction curve after allo-HCT for a patient whose eight pre-transplant factors are as follows: age, 50; DRI-R, high; PS, 1; HCT-CI, 1; conditioning intensity, MAC; HLA compatibility, matched; donor source, rPB; the number of allo-HCT, 1. This patient’s 1-year OS rate after allo-HCT is predicted at 59.1%. Users can also see the personalized 1-year PFS curve and cumulative incidence prediction curves of relapse/progression and NRM after allo-HCT. We provided an online interface to use this web application (https://predicted-os-after-transplantation.shinyapps.io/RSF_model/) and its source code (https://github.com/HiroshiOkamura-OCU/Prognosis_prediction_curves_in_allo-HCT).

**Figure 1.**
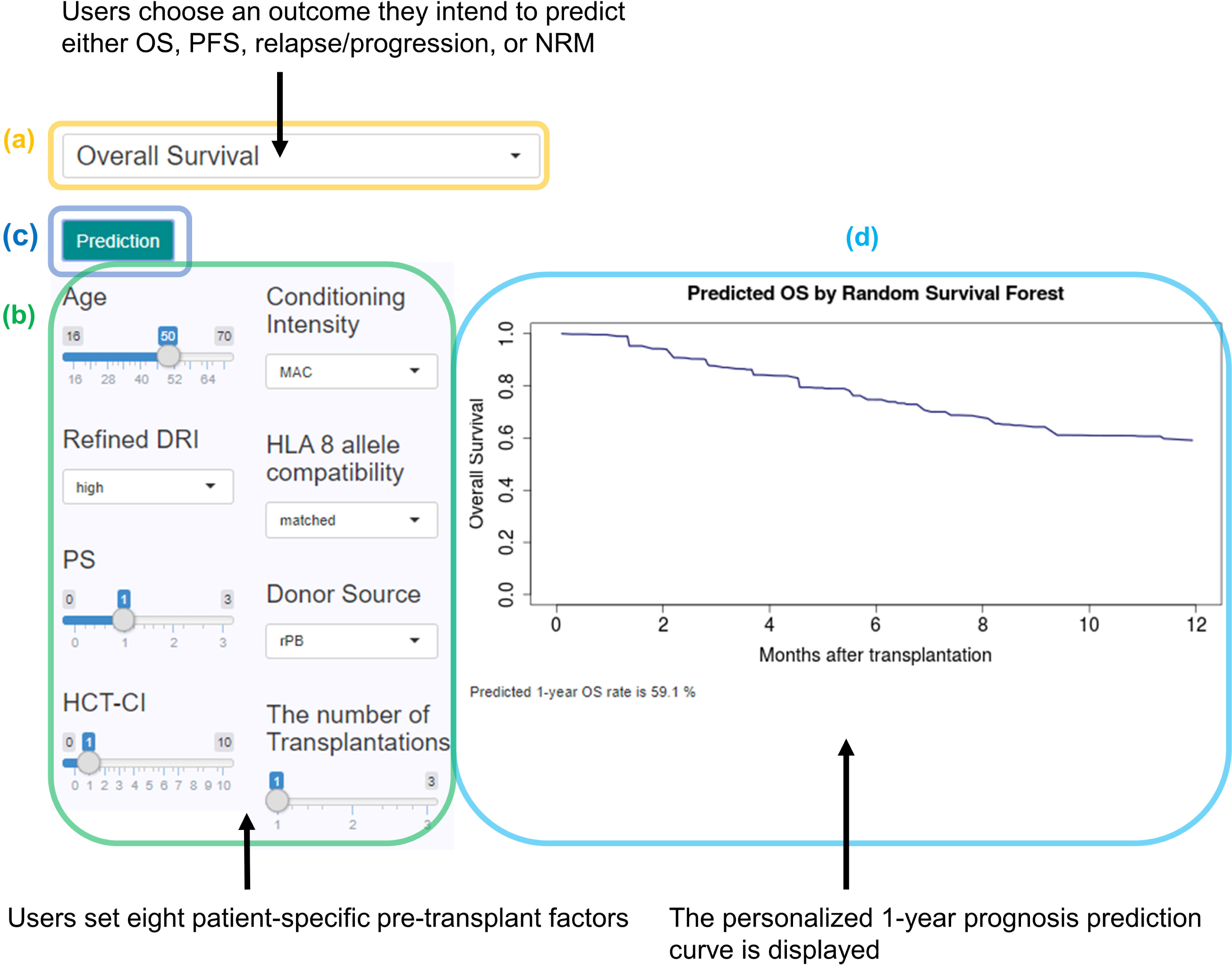
Interactive web application for plotting the personalized prognosis prediction curves after allo-HCT. The view and contents of the web application and an example of the personalized 1-year OS prediction curve which was displayed interactively for a patient (age, 50; DRI-R, high; PS, 1; HCT-CI, 1; conditioning intensity, MAC; HLA compatibility, matched; donor source, rPB; the number of allo-HCT, 1.) are shown. allo-HCT, allogeneic hematopoietic cell transplantation; DRI-R, refined Disease Risk Index; HCT-CI, Hematopoietic Cell Transplantation Comorbidity Index; HLA, human leukocyte antigen; MAC, myeloablative conditioning; NRM, non-relapse mortality; OS, overall survival; PFS, progression-free survival; PS, performance status; RIC, reduced intensity conditioning; rPB, related peripheral blood.

The predictive performances for each 1-year prognosis model in the RSF trained by the entire cohort is shown in Table S1. They were calculated using the 5-fold cross-validation method to prevent overfitting.

### Assessment of Predictive Performance

The AUCs calculated by the 5-fold cross-validation in the training cohort for 1-year OS, PFS, relapse/progression, and NRM per hyperparameter in the RSF model are shown in Table S2. The optimal number of variable candidates for splitting a node in the RSF model for 1-year OS, PFS, relapse/progression, and NRM were 7, 2, 2, and 5, respectively.

The AUCs for 1-year OS, PFS, relapse/progression, and NRM in the test cohort were 0.70, 0.72, 0.73, and 0.77, respectively (Table S3). Time-sequential AUCs plotted per month from 3 to 12 months after allo-HCT for OS, PFS, relapse/progression, and NRM in the test cohort are shown in Figure 2. The predictive performances of RSF for OS and PFS were comparable to those of the Cox PH model at any time-point. The predictive performances of RSF for every outcome were superior to those of DRI-R or HCT-CI alone consistently at any given time-point.

**Figure 2.**
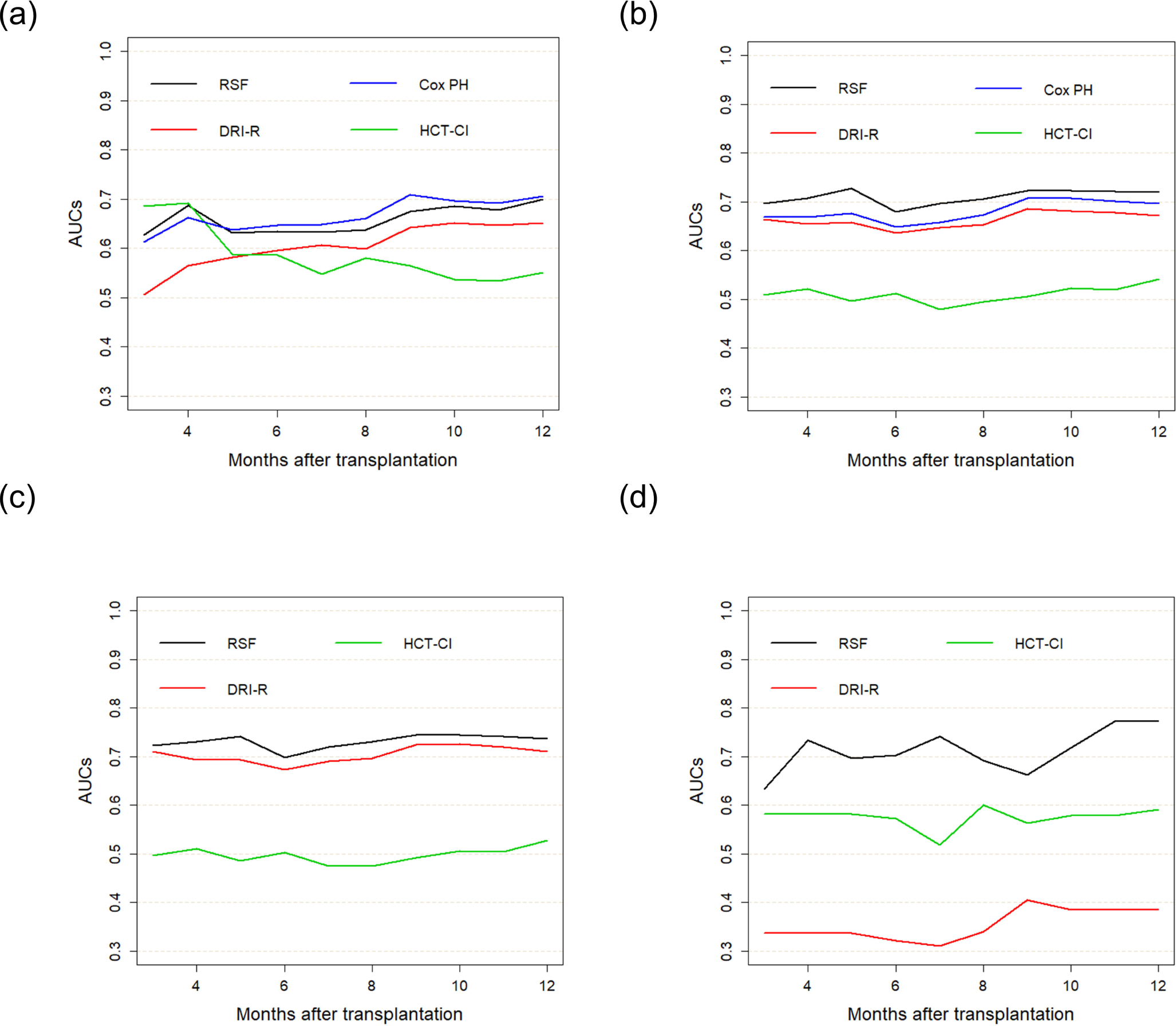
Time-sequential predictive performances. Time-sequential AUCs of RSF, Cox PH model, DRI-R, and HCT-CI for OS (a) and PFS (b) and those of RSF, DRI-R, and HCT-CI for relapse/progression (c) and NRM (d) are plotted per month from 3 to 12 months after allo-HCT. allo-HCT, allogeneic hematopoietic cell transplantation; AUCs; area under the receiver-operating characteristic curves; Cox PH, Cox proportional hazard; DRI-R, refined Disease Risk Index; HCT-CI, Hematopoietic Cell Transplantation Comorbidity Index; NRM, non-relapse mortality; OS, overall survival; PFS, progression-free survival; RSF, random survival forest.

## DISCUSSION

We succeeded in developing an interactive web application tool with GUI capable of accurately, objectively, and rapidly generating the four types of personalized 1-year prognosis prediction curves after allo-HCT in just a few steps using machine learning. To the best of our knowledge, there have been no previous reports regarding the development of a similar interactive tool that can provide various types of patient-specific prognosis prediction curves in allo-HCT.

Some predictive indices of prognosis after allo-HCT such as DRI-R, HCT-CI, EBMT risk score, and pre-transplantation assessment of mortality score have been reported.^2, 6–8, 20^ Although these indices can stratify prognosis after allo-HCT, transplant clinicians need to empirically consider various additional factors, such as age, PS, donor source, or conditioning regimen, to estimate patient-specific prognosis in clinical practice. Our web application tool can calculate and plot personalized prognosis prediction curves more objectively than a clinician’s empirical estimate and more accurately than any single prognosis index.

Shouval et al. developed a website to predict prognosis after allo-HCT using the alternating decision tree (ADTree), which is a machine learning model.^21^ Users can obtain personalized prognostic predictive information such as day 100 mortality by providing certain individual pre-transplant factors for a new transplant candidate. However, the ADTree model cannot treat time-to-event data, whereas the RSF is a machine learning algorithm for survival analysis that can take the right-censored data or competing risks into consideration.^12, 22^ Hence, our web application can not only provide prognosis prediction value for a fixed time point but also plot continuous survival and cumulative incidence prediction curves until 1 year after allo-HCT (Figure 1). Moreover, users can simulate various transplant settings for a particular patient intuitively with just a few clicks using mobile devices such as tablets and smartphones as well as computers because our web application was developed using shiny/R library to ensure interactivity. Consequently, our web application can help transplant clinicians inform a new allo-HCT candidate of their personalized prognosis estimate in clinical practice, and it can provide both the clinician and candidate with valuable information to help make clinical decisions.

Shouval et al. also reported on the prognosis prediction performance after allo-HCT by ADTree using registry data in European Group for Blood and Marrow Transplantation (EBMT) and Italian Group for Bone Marrow Transplantation (GITMO) and showed that the time-dependent AUCs for 2-year OS were 0.66 and 0.65, respectively.^21, 23^ Their prediction model developed using large amounts of patient data is quite relevant, because it can be used widely as a generalized prediction model in allo-HCT fields. However, various biases related to transplant institutes or countries are inherent in allo-HCT. For example, there are differences in the indications for allo-HCT, transplant experiences, patient characteristics, and strategy for donor source selection, conditioning regimen, or transplant-related complications among the transplant institutes or countries. Therefore, some reports have shown that there is a significant difference in prognosis after allo-HCT among transplant centers or countries.^24, 25^ The prediction model developed using data from a single institution is protected from bias based on the differences among the transplant institutes as long as it is used within that institute, because the patient characteristics or transplant procedures similar to those of the training cohort by which the prediction model was developed are likely to be inherited by future patients at that institute. Furthermore, the noise of data collected from a single institute may be less than that of multicenter data, because the methods of clinical assessment or data collection could be consistent in a single institute. Hence, an in-house developed prognosis prediction model may have more advantages for prognosis predictive performance after allo-HCT in a particular institute than the prediction model based on data sourced from multiple institutions. Supporting this idea, Fuse et al. also showed that the predictive performance of relapse in a single institute where the predictive model was developed using ADTree was better than that in another institute using the identical predictive model (AUC; 0.75 vs 0.67).^26^ We have published the source code of our web application on the Internet. Each transplant center can also develop a web application using an in-house prognosis prediction model with its past patient data.

To assess the predictive performance, we developed predictive models with the training cohort, which is the former group generated by splitting the entire cohort into two groups time-sequentially, and their predictive performances were assessed in the test cohort, the latter group. This assessment method accords with the Type 2b of prediction model studies covered by the TRIPOD statement.^16^ As a result, the predictive performances for 1-year OS, PFS, relapse/progression in the test cohort were worse than those in the training cohort (Tables S2 and S3). These results could be attributed to the difference in the year of allo-HCT between the training and test cohorts. The predictive performances of the RSF model trained by the entire cohort, which was used in the web application, is shown in Table S1. However, the predictive performances for the new allo-HCT candidates in this web application may not be as high as those shown in Table S1 because the allo-HCT procedures and patient characteristics can change over time.

This study has several limitations. First, the predictive performance of our web application for new allo-HCT candidates in other institutes is unknown since our model has not obtained external validation. However, as mentioned above, we suppose it would be suitable for a transplant center, which contains many allo-HCT data, to develop an in-house model for more accurate prognosis prediction. Therefore, we provide the source codes, which will help each transplant institute to develop an in-house web application for personalized prognosis prediction after allo-HCT and assess its predictive performance. Second, as a retrospective analysis, this web application may be susceptible to selection bias, which may limit the interpretation of the prediction results. Third, we restricted the number of pre-transplant predictors to eight to ensure the usability of this web application tool. These refined eight pre-transplant parameters could be easily and non-invasively obtained at a low cost. However, there may be other combinations of pre-transplant factors that are better for prognosis prediction. Therefore, further studies are required.

In conclusion, we developed an interactive web application tool capable of plotting four kinds of personalized prognosis prediction curves after allo-HCT objectively through the integration of eight pre-transplant factors using RSF, and confirmed their promising predictive performances. The predictive information obtained by this web application could allow transplant clinicians to inform a new allo-HCT candidate of their objective personalized prognosis prediction in clinical practice and could support the clinician’s and the candidate’s decision-making.

## Data Availability

The datasets generated during and/or analyzed during the current study are not publicly available due to the ethically sensitive nature of the research. Although they may be provided by the corresponding author on reasonable request, its request must be submitted to the Research Ethics Committee in our hospital and be approved.

## Acknowledgments

We wish to thank the Hematopoietic Cell Transplant Coordinator of Osaka City University Hospital, Ms. Umemoto and Ms. Tanaka for assistance with data entry.

## ABBEREVIATIONS

ADTree: alternating decision tree
allo-HCT: allogeneic hematopoietic cell transplantation
AUC: area under the receiver-operating characteristic curve
CB: cord blood
CI: confidence interval
Cox PH: Cox proportional hazard
DRI-R: refined disease risk index
EBMT: European Group for Blood and Marrow Transplantation
GITMO: Italian Group for Bone Marrow Transplantation
GUI: graphical user interface
haplo-PB: haploidentical peripheral blood
HCT-CI: hematopoietic cell transplantation comorbidity index
HLA: human leukocyte antigen
MAC: myeloablative conditioning
NRM: non-relapse mortality
OS: overall survival
PFS: progression-free survival
PS: performance status
rBM: related bone marrow
RIC: reduced intensity conditioning
rPB: related peripheral blood
RSF: random survival forest
uBM: unrelated bone marrow

## Notes

**Disclosure:** The authors declare no conflicts of interest.

**Funding:** This work was supported by the Japan Society for the Promotion of Science (JSPS) KAKENHI Grant (number 17K09017).

### Competing Interest Statement

The authors have declared no competing interest.

### Funding Statement

This work was supported by the Japan Society for the Promotion of Science (JSPS) KAKENHI Grant (number 17K09017).

